# Association of coffee consumption in metabolic syndrome: a cross-sectional and Mendelian randomization study in UK Biobank

**DOI:** 10.1101/2023.08.11.23293897

**Authors:** Tommy Hon Ting Wong, Shan Luo, Shiu Lun Au Yeung, Jimmy Chun Yu Louie

**Author notes:** Corresponding author: Dr. Jimmy Chun Yu Louie Address: Department of Nursing and Allied Health, School of Health Sciences, Swinburne University of Technology, Melbourne, Australia.

## Abstract

**Objective:** To investigate the associations between coffee consumption and metabolic syndrome and its component conditions, as well as the effect of using milk, sugar, and artificial sweetener on these associations.

**Research Design and Methods:** The cross-sectional analysis included 351,805 participants from the UK Biobank. Coffee consumption data was obtained using food frequency questionnaire and 24-hour recall and metabolic syndrome was ascertained based on blood biochemistry results and self-reported medication use. Odds ratios were calculated using multivariable logistic regression adjusted for lifestyle and socioeconomic factors, with verification using two-sample Mendelian randomization.

**Results:** Coffee consumption up to 2 cups per day was inversely associated with metabolic syndrome (1 cup/day, OR: 0.88, 95% CI: 0.85, 0.92; 2 cups/day, OR: 0.90, 95% CI: 0.86, 0.93) while associations at higher intakes were near null. Mendelian randomization did not support a causal association between coffee intake and metabolic syndrome. Both self-reported and genetically predicted high coffee consumption (4 cups per day or more) were associated with central obesity. The inverse association between coffee consumption and metabolic syndrome was more profound among drinkers of ground coffee than those of instant coffee. Results were similar when stratified by the use of milk and sugar, yet the use of artificial sweetener with coffee was positively associated with metabolic syndrome and all component conditions.

**Conclusions:** Coffee consumption likely increase the risk of central obesity but unlikely impact risk of metabolic syndrome. The possible health effect of using artificial sweetener with coffee warrant investigations in future studies.

## Introduction

Metabolic syndrome is a condition that involves the co-occurrence of multiple metabolic abnormalities, including high fasting blood glucose level, dyslipidemia, high blood pressure, and central obesity (1). Patients meeting the criteria of metabolic syndrome had higher risks of developing type 2 diabetes (2) and cardiovascular diseases (3). Given metabolic syndrome was estimated to be affecting 20 – 30% of the global population (4–7) and is a known risk factor for type 2 diabetes (2) and cardiovascular diseases (3), identifying modifiable factors contributing to increased risk of metabolic syndrome would be of great public health interest.

A recent systematic review of cross-sectional studies suggested an inverse relationship between coffee consumption and metabolic syndrome (8). Possible mechanisms may include an improvement in energy balance which was contributed by caffeine intake (9), as well as antioxidative and anti-inflammatory effects associated with polyphenols in coffee (10). However, our recent meta-analysis (8) found that coffee consumption was not longitudinally associated with metabolic syndrome. These are also consistent with randomized controlled trials (RCTs) showing habitual coffee consumption did not affect blood glucose and lipid levels (11; 12), which indirectly suggest previous observational studies may be confounded by sociodemographic factors or reverse causation. Nevertheless, it remains possible that the effect of coffee with metabolic syndrome may differ by coffee subtypes or how the coffee was being prepared (e.g. the use of milk and sweetener).

The primary aim of this study is to assess the cross-sectional association of overall coffee consumption in metabolic syndrome and its components (i.e., high fasting blood glucose, high triglycerides, central obesity, high blood pressure, and low high-density-lipoprotein-cholesterol (HDL-cholesterol) using a large population-based study. We also conducted a Mendelian randomization study, which used genetic variants allocated at conception and hence less vulnerable to confounding, to verify the observational association (13). The secondary aims were to investigate the association of coffee consumption in metabolic syndrome differed by coffee types and the use of milk, sugar, and artificial sweetener.

## Research Design and Methods

### The UK Biobank

The UK Biobank is a large-scale prospective cohort aiming to improve the understanding of the prevention, diagnosis, and treatment of a wide range of common and life-threatening diseases in middle to old-aged people. The design and the protocol of this project has been previously published (14; 15). In brief, approximately 500,000 participants with 40 – 69 years of age living in the United Kingdom were recruited during 2006 – 2010. Data regarding demographics, socioeconomic status, health, and diet were collected in 22 assessment centers across the United Kingdom and blood samples were also taken for biochemical analyses. All UK Biobank participants were genotyped using either the Affymetrix UK BiLEVE Axiom or Affymetrix UK Biobank Axiom array. The UK Biobank project was approved by the National Information Governance Board for Health and Social Care and the NHS North West Multicenter Research Ethics Committee (reference number: 11/NW/0382). Written consent was obtained from all participants.

### Habitual coffee intake

Participants were asked “How many cups of coffee do you drink each day (include decaffeinated coffee)?” and they were prompted to enter their average amount of consumption over the last year in positive integers in a food frequency questionnaire (FFQ). Participants were also asked to select the type of coffee that they used to drink at that time and available responses included “decaffeinated coffee”, “instant coffee”, “ground coffee (include espresso, filter etc.)”, and “other type of coffee”. Participants recruited during 2009-2010 also filled in a detailed 24-hour recall dietary questionnaire in addition to the FFQ. In the 24-hr recall questionnaire they were asked “Did you drink any coffee yesterday?” with a “Yes/No” option. For those who selected “Yes” they were further asked for their consumption (in cups or mugs) of different types of coffee on the previous day. Available options for coffee types included instant coffee, filtered coffee, cappuccino, latte, espresso, decaffeinated coffee, and other coffee types. Participants also reported whether they added milk to instant and filtered coffee, as well as sugar or artificial sweetener (in teaspoon) when drinking coffee in general.

### Outcomes

The primary outcome was metabolic syndrome, which was defined by the International Diabetes Federation in 2009 (1) as having any 3 of the following: high fasting glucose, high triglycerides, central obesity, high blood pressure, and low HDL-cholesterol (secondary outcomes). The detailed criteria for each condition was listed in **Supplemental Table 1**. Waist circumference was measured in cm using a measurement tape (SECA). Blood pressure (mmHg) was measured twice using the IntelliSense blood pressure monitor model HEM-907XL (Omron) for each participant and the average of the two measurements were used in this study. Serum levels of glucose (mmol/L), triglycerides (mmol/L), and HDL-cholesterol (mmol/L) were all measured using the Beckman Coulter AU5800 platform. Medications for treating abnormal levels of glucose, lipids, and blood pressure were self-reported and the corresponding medication code were shown in **Supplemental Table 2**. The details of case and control ascertainment and handling of missing data was stated in **Supplemental Information**.

### Potential confounders

We considered age, sex, socioeconomic position (Townsend Deprivation index, and highest qualification obtained), lifestyle factors (smoking status, alcohol intake frequency, physical activity, and intakes of fruit, vegetable, and tea) as possible confounders. Details of measurement were provided in the **Supplemental Information**.

### Genetic predictors of coffee consumption

Genetic predictors of coffee consumption, i.e., single nucleotide polymorphisms (SNPs), were retrieved from a genome-wide association study (GWAS) of predominantly European ancestry in the Coffee and Caffeine Genetics Consortium (CCGC) (16). These included (i) coffee consumption in cups per day among coffee drinkers (phenotype 1, *n* = 129,488) and (ii) no/low versus high coffee consumption (phenotype 2, *n* = 65,842). Only independent SNPs (*r^2^* < 0.001) that were genome-wide significant (*p <* 5 × 10^-8^) in the trans-ethnic meta-analysis were selected.

### Genetic associations of the outcomes

The genetic associations between the SNPs identified for cups of coffee consumed (phenotype 1) and being a coffee drinker (phenotype 2) with metabolic syndromes and its components were obtained from the UK Biobank participants of European ancestry. The genetic associations of outcomes were obtained using multivariable logistic regression, adjusted for age, sex, top 20 principal components, genotype array, and assessment center. Participants with poor quality genotyping (missing rate > 0.01), ≥ 10 putative third-degree relatives, with sex chromosome aneuploidy, with a mismatch between genetic sex and reported sex, and those with a mismatch in reported ethnicity and genetic ethnicity were excluded. Up to 360,903 participants provided data for the genetic associations of outcomes.

## Statistical analyses

### Cross sectional analyses

We assessed the cross-sectional associations of coffee consumption in metabolic syndrome and its components using multivariable logistic regressions with zero consumers as the reference group. Effect estimates were adjusted for potential confounders (age, sex, Townsend deprivation index, highest qualification attained, smoking status, alcohol intake frequency, physical activity level, fruit intake, vegetable intake, and tea consumption). We also included coffee type as a covariate when analyzing data collected from 24-hour recalls. To investigate the association between using milk, sugar, and artificial sweetener with coffee and all outcomes, we stratified the main analyses by these factors whenever possible. Stratification by milk use was carried out for instant and filtered coffee, while stratification by use of sugar and artificial sweetener was also carried out for all coffee types except espresso due to a lack of cases.

### Mendelian randomization analysis

We calculated the *R^2^* (only phenotype 1) and *F* statistics of the instruments as an indicator of instrument strength based on equations used in a previous Mendelian randomization study (17), where a higher F statistic indicated lower evidence for weak instrument bias. Effect alleles of all instruments were aligned to the coffee consumption-increasing allele. We assessed the causal association of coffee consumption in metabolic syndrome and its component conditions using inverse variance weighted (IVW) with multiplicative random effect models. The IVW estimator assumes balanced horizontal pleiotropy (18) and high heterogeneity among instrument-specific Wald Ratios based on Cochran’s *Q* test may imply the inclusion of invalid instruments. To assess the robustness of our findings, we also conducted a range of sensitivity analyses which were based on different assumptions. The MR-Egger estimator allows for overall directional pleiotropic effects by assuming the instrument strength is independent of the direct effect (InSIDE assumption). An MR-Egger intercept with a *p*-value < 0.05 reflects the presence of horizontal pleiotropic effect (18). The weighted median method assumes at least 50% of the weight come from valid SNPs (19). Since smoking status and alcohol consumption were well-recognized confounding factors in the associations between coffee consumption and various health outcomes (20) and genetic predictors of coffee consumption were found to be associated with both traits as well (20) we conducted multivariable MR to obtain effect estimates adjusted for alcohol consumption and liability to smoking initiation. Details regarding the multivariable Mendelian randomization analysis was provided in the **Supplemental Information**.

To correct for multiple testing, the *p*-value threshold for statistical significance for all analyses was set at 0.0083 (0.05 divided by 6 outcomes).

## Results

### Participant characteristics

Participants who had withdrawn, were not British, without coffee consumption data, and without data on any confounding variables were excluded from all analyses. Those with missing individual outcome variables were excluded from the corresponding analyses. The detail exclusion flowchart was shown in **Figure 1**. The characteristics of all included participants (*n* = 351,805) were shown in **Table 1**. Participants who consumed coffee were more likely to be male, more likely to be current smoker, consumed alcohol more frequently, had higher education attainment, and drank less tea.

**Figure 1.**
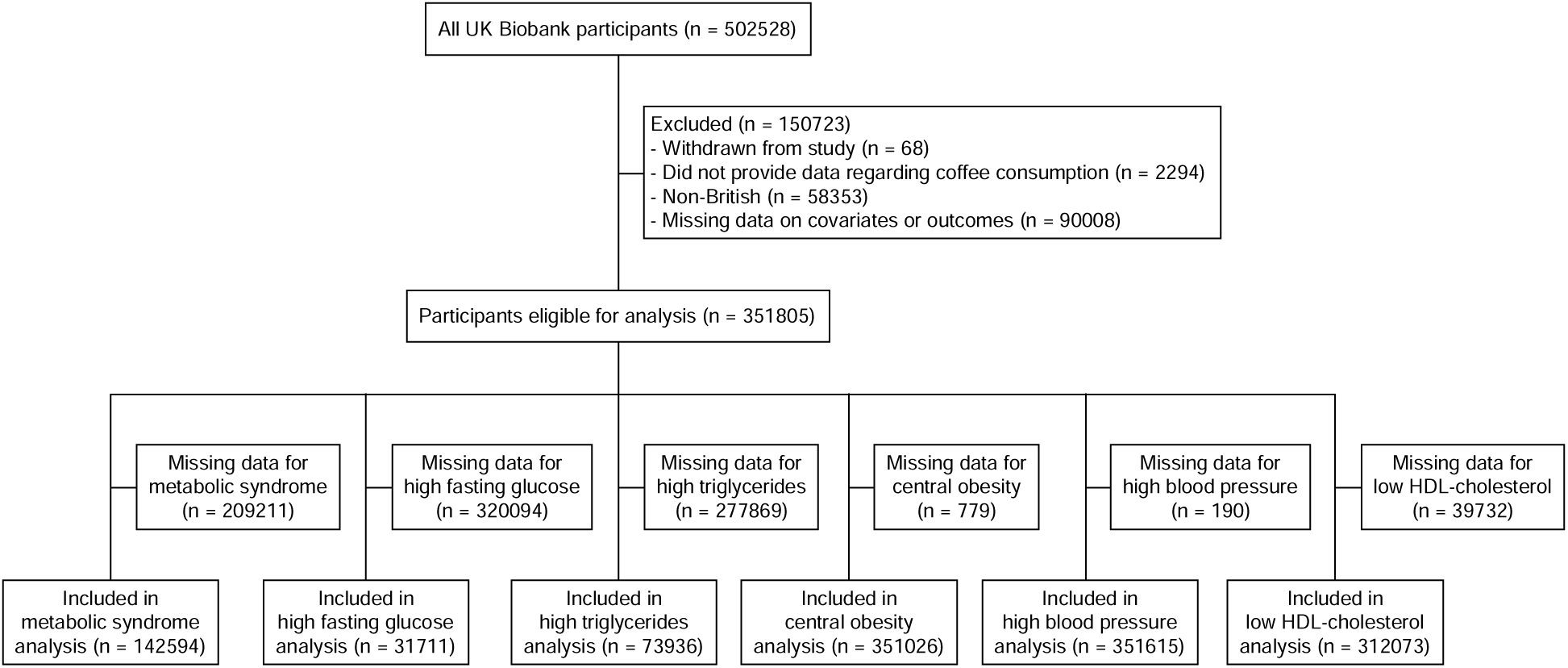
Participant exclusion flowchart for the cross-sectional analysis.

**Table 1.** characteristics of participants included in cross-sectional analysis (n = 351,805)

| Variables | Coffee consumption (cups/day) |  |  |  |  |  |  |  |
| --- | --- | --- | --- | --- | --- | --- | --- | --- |
|  | 0 | <1 | 1 | 2 | 3 | 4 | 5 | 6 or more |
| Participants, n | 72860 | 24309 | 71116 | 67516 | 44629 | 30940 | 18260 | 22175 |
| Age, years | 55.5 (8.2) | 56.4 (7.9) | 57.3 (8.0) | 57.2 (7.9) | 57.0 (7.9) | 56.6 (7.9) | 56.0 (8.0) | 55.6 (8.0) |
| Male, % | 43.5 | 47.0 | 45.2 | 48.1 | 49.9 | 52.2 | 54.2 | 55.6 |
| Smoking status, % |  |  |  |  |  |  |  |  |
| Never | 57.6 | 58.3 | 57.4 | 55.7 | 54.3 | 50.9 | 47.2 | 39.9 |
| Previous | 33.2 | 34.2 | 35.7 | 36.1 | 36.6 | 37.4 | 37.2 | 35.9 |
| Current | 9.2 | 7.5 | 6.9 | 8.2 | 9.0 | 11.7 | 15.6 | 24.1 |
| Alcohol consumption frequency, % |  |  |  |  |  |  |  |  |
| Monthly or less | 37.7 | 27.1 | 24.9 | 21.8 | 21.6 | 22.7 | 25.8 | 32.3 |
| Weekly | 46.0 | 52.7 | 53.1 | 53.2 | 52.5 | 52.4 | 51.1 | 46.9 |
| Daily | 16.3 | 20.2 | 22.0 | 25.0 | 25.8 | 24.8 | 23.1 | 20.8 |
| Fruit intake, tbsp | 3.0 (2.5) | 3.1 (2.4) | 3.2 (2.5) | 3.2 (2.5) | 3.0 (2.4) | 2.9 (2.3) | 2.8 (2.4) | 2.7 (2.6) |
| Vegetable intake, tbsp | 4.7 (3.3) | 4.7 (3.0) | 4.9 (3.0) | 4.9 (3.0) | 4.8 (3.0) | 4.8 (3.1) | 4.7 (3.1) | 4.8 (3.5) |
| Tea intake, cups/day | 4.7 (3.3) | 4.6 (2.8) | 4.0 (2.5) | 3.3 (2.4) | 2.6 (2.2) | 2.2 (2.3) | 1.8 (2.5) | 1.9 (3.3) |
| IPAQ category, % |  |  |  |  |  |  |  |  |
| Low | 20.0 | 19.3 | 17.0 | 17.1 | 18.3 | 19.2 | 21.4 | 22.3 |
| Moderate | 39.0 | 42.0 | 41.6 | 42.1 | 42.1 | 41.3 | 39.6 | 38.1 |
| High | 41.0 | 38.7 | 41.4 | 40.8 | 39.6 | 39.5 | 39.0 | 39.6 |
| Highest qualification obtained, % |  |  |  |  |  |  |  |  |
| Below GCSE | 26.1 | 18.2 | 21.3 | 18.6 | 18.1 | 20.2 | 20.6 | 23.9 |
| GCSE | 24.7 | 21.8 | 21.5 | 21.3 | 20.9 | 22.0 | 22.8 | 24.1 |
| Advanced level | 7.7 | 8.4 | 7.6 | 7.7 | 7.7 | 7.4 | 7.3 | 7.6 |
| Degree holder | 41.6 | 51.6 | 49.7 | 52.4 | 53.3 | 50.4 | 49.4 | 44.5 |

Values are mean (SD) for age and percentages for count variables. Coffee consumption was analyzed as a categorical variable. GCSE, general certificate of secondary education. IPAQ, International Physical Activity Questionnaire.

### Association of coffee consumption and metabolic syndrome, and its components

Figure 2 shows the association between consuming cups of coffee per day and metabolic syndrome amongst 142,594 participants, after controlling for potential confounders. Participants consuming up to 3 cups of coffee per day were less likely to have metabolic syndrome when compared with coffee abstainers (<1 cup per day, OR: 0.91, 95% CI: 0.87, 0.96; 1 cup per day, OR: 0.88, 95% CI: 0.85, 0.92; 2 cups per day, OR: 0.90, 95% CI: 0.86, 0.93; 3 cups per day, OR: 0.94, 95% CI: 0.90, 0.99). However, consuming 4 cups of coffee or more per day was potentially associated with higher odds of having metabolic syndrome (4 cups per day, OR: 1.03, 95% CI: 0.98, 1.08; 5 cups per day, OR: 1.06, 95% CI: 0.99, 1.12; 6 cups per day, OR: 1.05, 95% CI: 0.99, 1.11). Coffee consumption was also positively associated with high fasting glucose and high triglycerides, but inversely associated with having high blood pressure and low HDL-cholesterol (Figure 2). The association between coffee consumption and central obesity was J-shaped.

**Figure 2.**
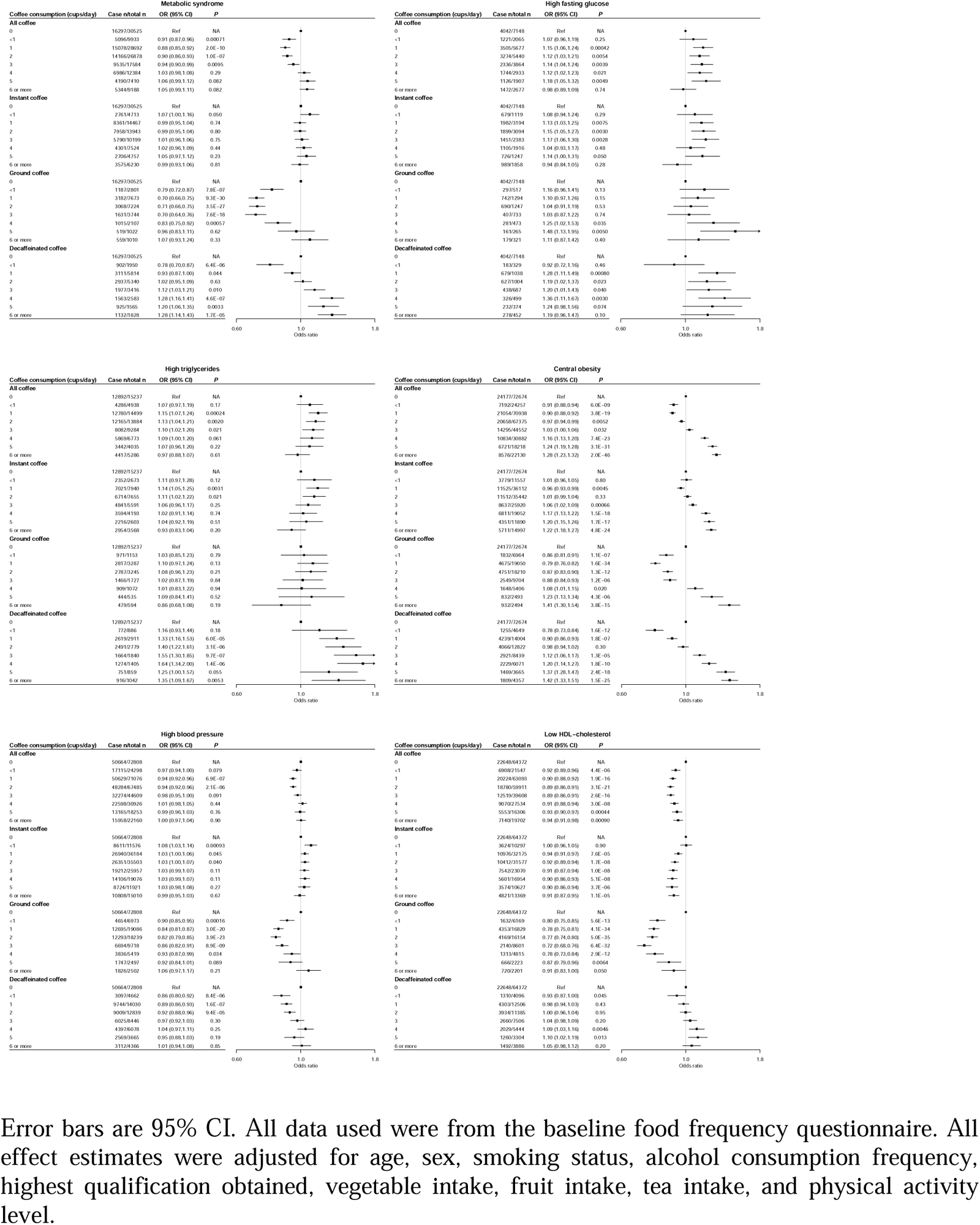
Associations between overall coffee consumption, as well as stratified by coffee types, and metabolic syndrome, high fasting glucose, high triglycerides, central obesity, high blood pressure, and low HDL-cholesterol.

Using Mendelian randomization (8 instruments for cups of coffee consumed: *R^2^*: 0.4%, F statistics: 56.1, and the range of F statistics for the 3 instruments for liability to high coffee consumption: 44.4 – 100.0) (**Supplemental Table 3)**, we did not identify any association of these two coffee phenotypes with risk of metabolic syndrome using IVW (Figure 3). Although the instruments were heterogenous based on Cochran’s Q, there was no strong evidence for overall horizontal pleiotropy based on MR-Egger intercept (**Supplemental Tables 4 and 5**). Adjusting for the liability to smoke initiation and alcohol consumption did not lead to material changes in the results (**Supplemental Tables 6 – 9**). However, both coffee phenotypes were associated with higher risk of central obesity, with directionally consistent findings from the sensitivity analyses (Figure 3). After adjusting for genetic liabilities to smoking initiation and alcohol consumption, results of both phenotypes were similar to those of the univariate MR. There was suggestive evidence that both phenotypes were associated with higher risk of high fasting glucose, but not high triglycerides, high blood pressure and low HDL-cholesterol (Figure 3).

**Figure 3.**
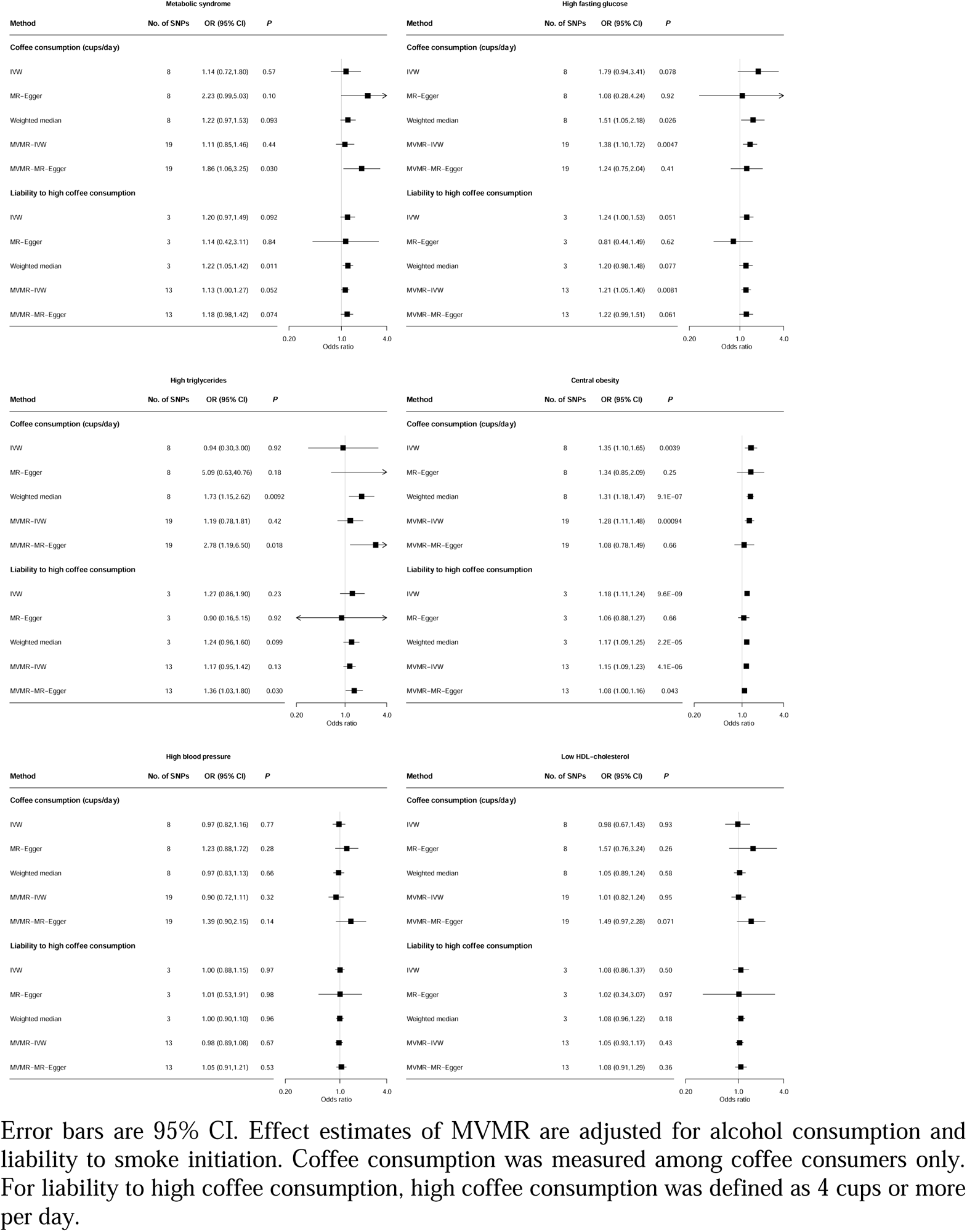
Results of MR analysis between coffee intake and metabolic syndrome, high fasting glucose, high triglycerides, central obesity, high blood pressure, and low HDL-cholesterol.

### Association of coffee subtypes, the use of sugar/milk, in metabolic syndrome and its components

The directions of associations observed among drinkers of instant coffee, ground coffee, and decaffeinated coffee mostly aligned with those of the main results with a few exceptions (Figure 2). For instance, null associations were observed between instant coffee intake with metabolic syndrome and high blood pressure, as well as between decaffeinated coffee intake and low HDL-cholesterol. Latte and cappuccino intake was inversely associated with metabolic syndrome and high blood pressure only, while all associations for espresso intake were near null (**Supplemental Figures 1 – 2**).

Results of analysis stratified by use of milk, sugar, and artificial sweetener were shown in **Supplemental Figures 1 – 4**. Stratifying by milk use did not affect any of the associations. Sugar use was inversely associated with high fasting glucose and central obesity among drinkers of instant coffee but not other coffee types.

Artificial sweetener use was positively associated with all outcomes among instant coffee consumers. Similarly, filtered coffee intake with artificial sweetener was positively associated with all outcomes except high fasting glucose and high triglycerides. Avoidance of artificial sweetener was inversely associated with metabolic syndrome, central obesity, and low HDL-cholesterol among filtered coffee drinkers, as well as metabolic syndrome and high blood pressure among latte and cappuccino drinkers.

## Discussion

In this large population-based study, we found habitual coffee consumption at 1-3 cups per day was associated with lower risk of metabolic syndrome. However, these associations were not replicated using Mendelian randomization, implying possible confounding in the observational analyses. We also observed that only ground and decaffeinated coffee intakes were inversely associated with metabolic syndrome, while a null association was observed among instant coffee drinkers. Moreover, we showed for the first time in Europeans that while milk and sugar use did not substantially affect the association between coffee intake and metabolic syndrome, the use of artificial sweetener was positively associated with metabolic syndrome and all component conditions among both instant and filtered coffee drinkers.

Our study was consistent with a previous smaller Danish study which also showed an inverse observational association of coffee consumption in metabolic syndrome but not in Mendelian randomization (21). As such, the inverse association observed in previous studies (22; 23) could possibly be due to residual confounding, such as variation in diet quality (24). However, we also observed a positive association of coffee consumption in central obesity in the observational and genetic analyses, a finding that was in line with a previous one-sample Mendelian randomization study (25) that observed a positive association between coffee consumption and obesity, but inconsistent with previous observational studies (26). The bioactive components in coffee unlikely explained this association, since caffeine consumption may enhance lipolysis and food-induced thermogenesis (27), while evidence regarding the effect of polyphenols found in coffee on preventing obesity have been inconsistent (28). Furthermore, a recent longitudinal study observed a mild association between habitual coffee consumption and waist circumference reduction (29). One possible cause behind our findings could be psychological stress, which is a well-known factor of obesity (30). Evidence from observational studies supported a positive association between caffeine intake and psychological stress (31; 32). Although cortisol, which was an established marker for psychological stress, was found to be causally unrelated to waist-to-hip ratio and body-mass-index (33), the authors acknowledged that genetically predicted cortisol may not be a direct reflection of stress, which would induce other biological responses other than cortisol elevation, such as the release of inflammatory cytokines (34). Further studies are warranted to investigate if psychological stress is in the same causal pathway as coffee intake and central obesity.

One novel aspect of this study was to explore the role of additives in the relation of coffee consumption in metabolic syndrome, where we showed the use of artificial sweeteners were associated with metabolic syndrome and all component conditions. There were reports linking artificial sweetener use with increased caloric intake, worsened glycemic control, long-term weight gain, and metabolic syndrome (35). One possible mechanism behind our finding is that since sweet taste is treated by the brain as a signal of incoming caloric content, the lack of energy in the artificial sweetener would weaken this perception, leading to dysregulation in nutrient signaling and subsequent homeostasis (36). Another proposed mechanism is that artificial sweetener intake could alter the microbiota composition in human gut, thereby inducing glucose intolerance (37). Nonetheless, while the evidence regarding the effect of artificial sweetener use on weight management and glycemic control was highly mixed (38; 39), our findings could be also explained by reverse causation where they used artificial sweeteners because of their poor metabolic health. Further longitudinal studies are needed to determine if the use of sugar and artificial sweeteners augments the association between coffee consumption and metabolic syndrome.

Strengths of this study include large sample size and the use of different approaches to interrogate the role of coffee consumption in metabolic syndrome, as well as the role of additives. Nevertheless, several limitations of this analysis should be acknowledged. First, the definitions of metabolic syndrome used in this study required fasting measurement of biomarkers, thus a portion of the participants had to be excluded from the analyses of certain outcomes and the results may possibly be affected by selection bias. Second, some analyses that were stratified by use of sugar and artificial sweetener were left with very few cases, thus the statistical power of such tests could be limited. Third, although we corrected for possible horizontal pleiotropy via alcohol and tobacco use using multivariable Mendelian randomization, the genetic instruments used in this analysis were also associated with tea consumption, as shown in a previous study (20), and hence may be biased by horizontal pleiotropy although the findings were generally consistent in other sensitivity analyses which were more robust to violation via horizontal pleiotropy, such as MR-Egger. Fourth, the use of a dichotomized exposure variable (phenotype 2) could possibly violate the instrumental variable assumption of MR (40). Finally, results from this study may not be generalizable to populations of non-European ancestry because the data used in this study were collected from participants of European origin only.

To conclude, low-to-moderate coffee intake (1-3 cups per day) was associated with lower risk of metabolic syndrome in cross-sectional analyses, but such associations were not supported by Mendelian randomization. However, high coffee consumption may contribute to higher risk of central obesity. Our findings also highlighted the variation in results due to coffee types and use of sugar and artificial sweeteners. Nonetheless, the findings should be verified in future longitudinal studies with data in coffee consumption and use of coffee additives, while the causal relationship between coffee intake and metabolic health outcomes should be verified in future Mendelian randomization studies involving more genetic instruments for coffee consumption.

## Data availability

The data used in this study can be requested via application to UK Biobank (https://www.ukbiobank.ac.uk).

## Acknowledgement

The present study was conducted under UK Biobank application number 44407.

## Authors’ contributions

JCYL and TWHT designed research; SL, AYSL, and JCYL provided essential materials; TWHT analyzed the data, performed statistical analysis, and drafted the paper; all authors critically evaluated and amended the manuscript. TWHT and JCYL had primary responsibility for the final content. All authors read and approved the final manuscript.

