## Supplemental figures 1-4 for "Association of coffee consumption in metabolic syndrome: a cross-sectional and Mendelian randomization study in UK Biobank"

Supplemental figure 1 – Association between latte and cappuccino consumption and all outcomes, stratified by the use of milk, sugar, and artificial sweetener.

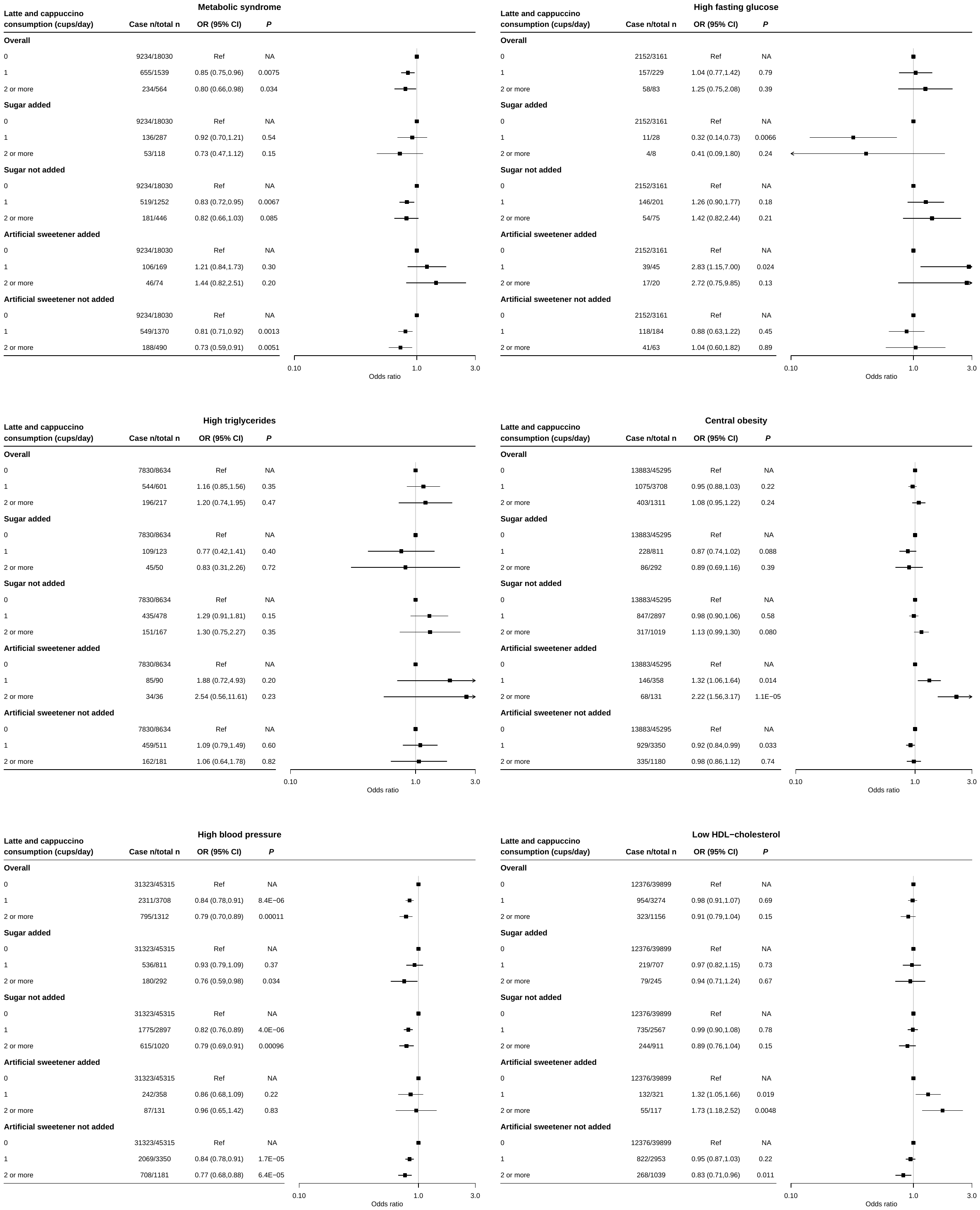

All data were from 24-hour recalls of UK Biobank. All effect estimates were adjusted for age, sex, smoking status, alcohol consumption frequency, vegetable intake, fruit intake, tea intake, physical activity level, and highest qualification obtained. Error bars depict 95% CI.

Supplemental figure 2 – Association between espresso consumption and all outcomes, stratified by the use of milk, sugar, and artificial sweetener.

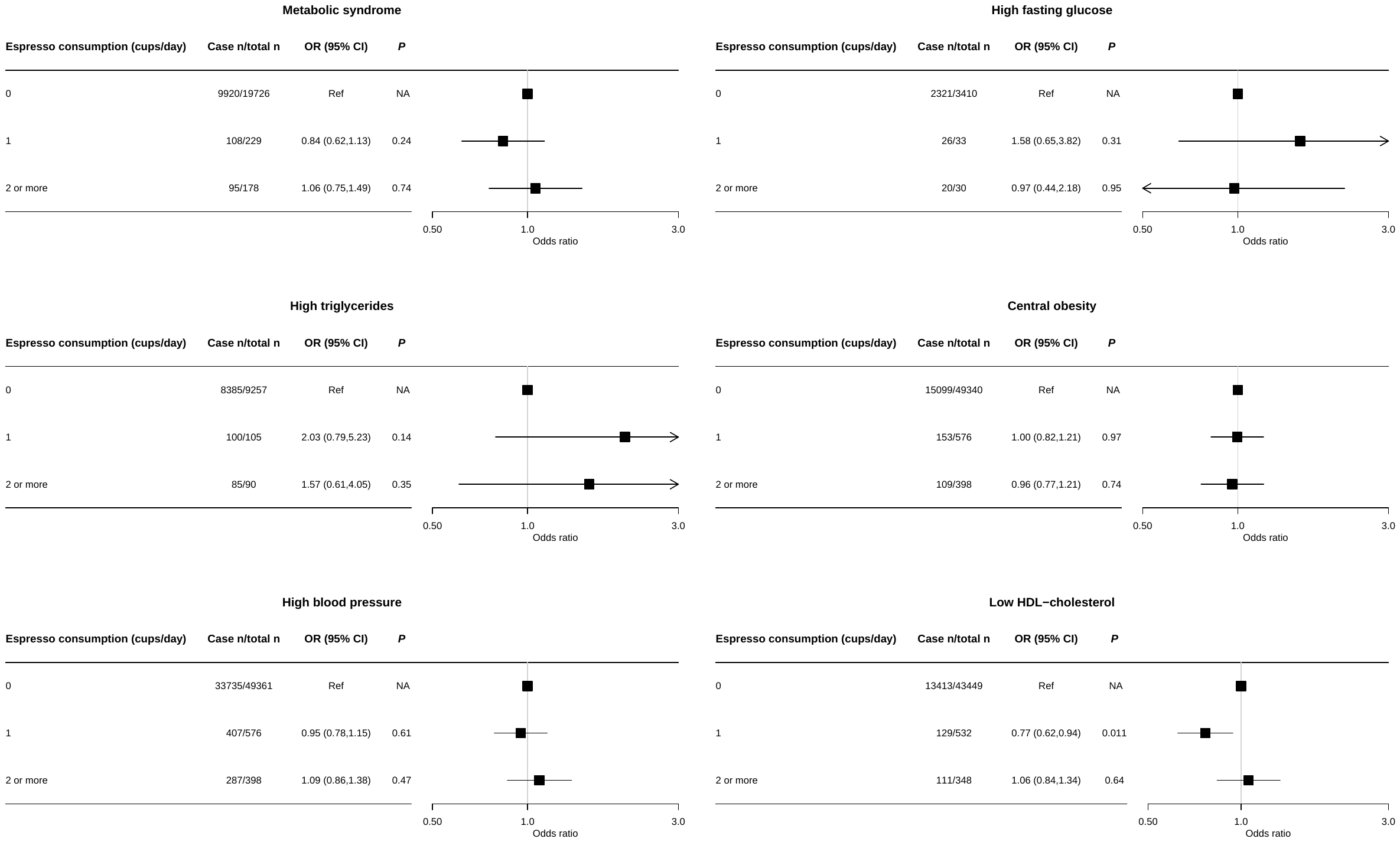

All data were from 24-hour recalls of UK Biobank. All effect estimates were adjusted for age, sex, smoking status, alcohol consumption frequency, vegetable intake, fruit intake, tea intake, physical activity level, and highest qualification obtained. Error bars depict 95% CI.

Supplemental figure 3 – Association between instant coffee consumption and all outcomes, stratified by the use of milk, sugar, and artificial sweetener.

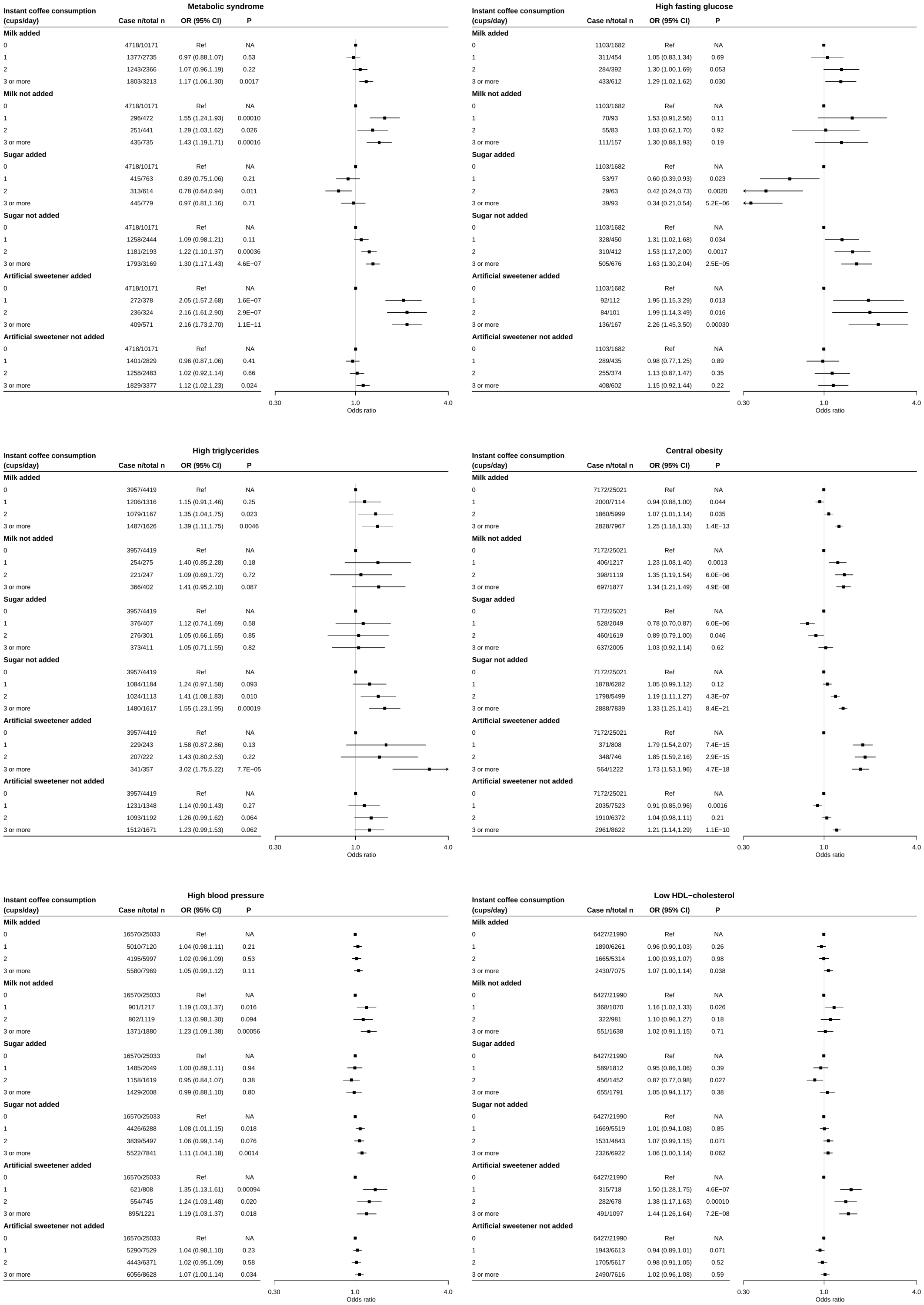

All data were from 24-hour recalls of UK Biobank. All effect estimates were adjusted for age, sex, smoking status, alcohol consumption frequency, vegetable intake, fruit intake, tea intake, physical activity level, and highest qualification obtained. Error bars depict 95% CI.

Supplemental figure 4 – Association between filtered coffee consumption and all outcomes, stratified by the use of milk, sugar, and artificial sweetener.

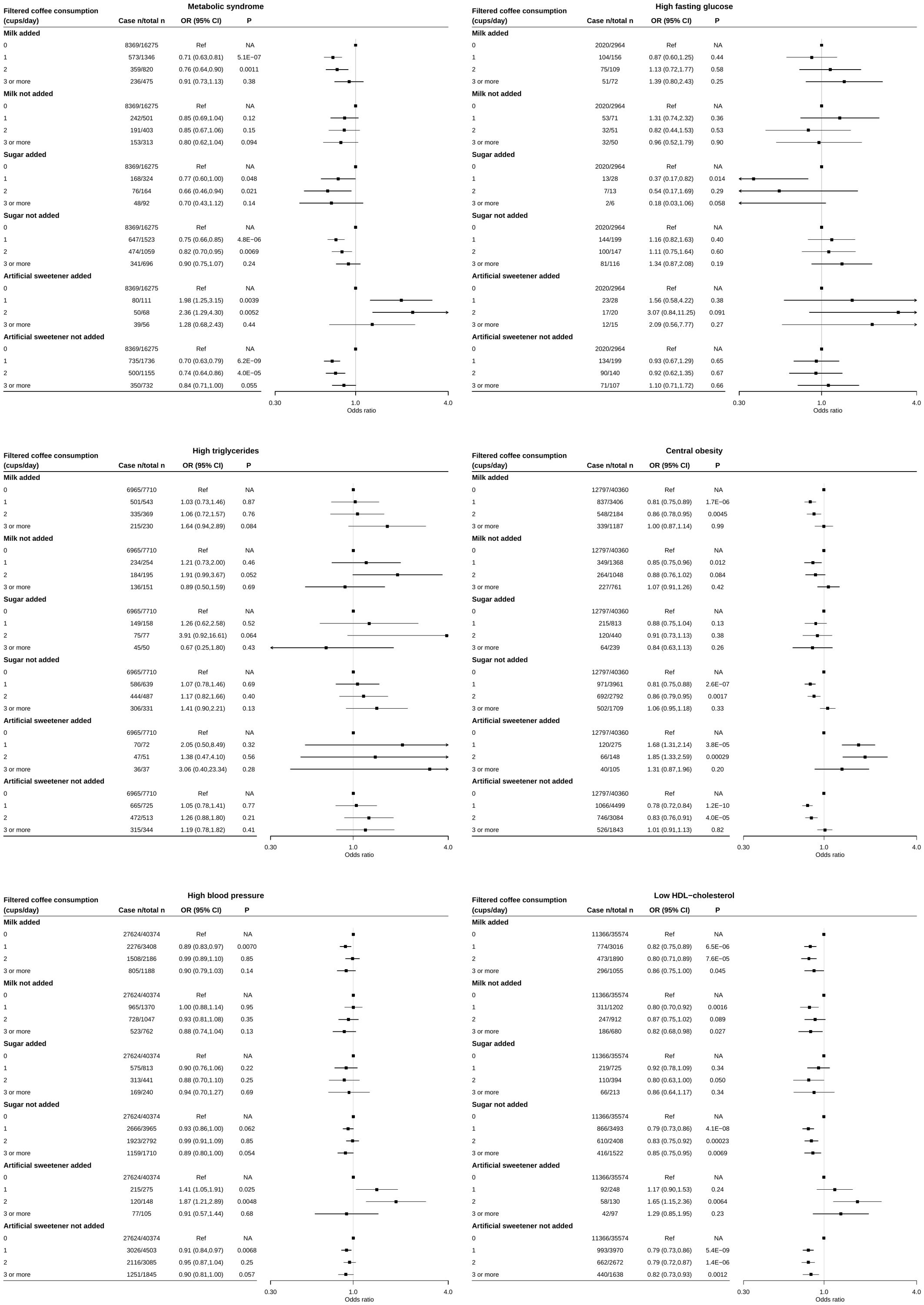

All data were from 24-hour recalls of UK Biobank. All effect estimates were adjusted for age, sex, smoking status, alcohol consumption frequency, vegetable intake, fruit intake, tea intake, physical activity level, and highest qualification obtained. Error bars depict 95% CI.
