## Supplemental material for "Association of coffee consumption in metabolic syndrome: a cross-sectional and Mendelian randomization study in UK Biobank"

### Supplemental information

#### Case and control ascertainment of outcomes

For each outcome, cases were defined as meeting the ascertainment criteria, while controls were defined as not meeting the ascertainment criteria without missing data. The handling of missing data was stated in the next section. For metabolic syndrome, cases were defined as meeting the criteria of cases in 3 or more of the component conditions. Controls were defined as meeting the criteria of controls in 3 or more of the component conditions. Those who did not meet the criteria for either case or control were excluded from the analysis.

#### Handling of missing data

Since the UK Biobank collected random blood samples during the baseline recruitment, only 6% of the total participants gave fasting blood samples, which was defined as blood samples given with at least 7 hours of fasting in this study. Therefore, for those who did not provide a fasting blood sample, a case of high fasting glucose level was defined as taking medication to control high fasting glucose level, while those not taking medication would be treated as missing as we could not be sure if their fasting levels would meet the criteria. The same logic was applied when ascertaining cases for high triglycerides.

For participants that did not provide data regarding medication use, a case of high fasting glucose levels was defined as having fasting glucose levels exceeding the threshold as stated in **Supplemental Table 2**, while the rest would be treated as missing as we could not be sure if the observed fasting glucose level was due to taking medication. The same logic applied when ascertaining cases for high triglycerides, high blood pressure, and low HDL-cholesterol levels.

#### Measurement of confounding variables

Socioeconomic position was depicted by the Townsend Deprivation index (1). Smoking status, alcohol intake frequency, and highest qualification obtained were self-reported and recoded as follows: smoking status (never/previous/current), alcohol intake frequency (monthly or less/weekly/daily), highest qualification obtained (below general certificate of secondary education (GCSE), GCSE, A-level, or degree holder). Physical activity level was measured using the validated International Physical Activity Questionnaire (IPAQ) and was classified as low, moderate, or high according to the guidelines laid down by the IPAQ research committee (2). Intakes of fruit, vegetable, and tea were self-reported by the participants. Fruit intake was calculated by summing the intake of fresh and dried fruit while vegetable intake was calculated by summing the cooked and raw vegetable intake. Tea intake was measured in cups per day.

#### Multivariable Mendelian Randomization

We identified SNPs that were associated with alcohol consumption and liabilities to smoking initiation with genome-wide significance (*p <* 5 × 10^-8^) from the GWAS & Sequencing Consortium of Alcohol and Nicotine use (GSCAN) (3) study. The GSCAN study consisted participants only of European ancestry, that were not overlap with participants in the UK Biobank cohort (*n* for smoking initiation = 249,171; *n* for alcohol consumption = 226,223). For multivariable MR, the genetic predictors of coffee consumption were based on the results from stage one of the CCGC meta-analysis (consisted of European participants only), as the full summary statistics of stage two analyses were not publicly available. We excluded SNPs that were in linkage disequilibrium with other instruments (*r*^2^ < 0.1) and aligned each genetic association for exposure and outcome on the same effect allele. Proxy SNPs (*r^2^* ≥ 0.8) were used if the target SNPs were not available from any of the datasets. Causal effect estimates of coffee consumption on metabolic syndrome and the individual components were calculated using multivariable IVW and multivariable MR-Egger (4; 5).

R packages used

All analyses were done using R version 4.1.1 (6). All MR analyses were done using the “TwoSampleMR” package (7; 8) and “MVMR” package (9).

2. IPAQ Research Committee.: Guidelines for Data Processing and Analysis of the International Physical Activity Questionnaire (IPAQ) - Short and Long Forms. 2005

3. Liu M, Jiang Y, Wedow R, Li Y, Brazel DM, Chen F, Datta G, Davila-Velderrain J, McGuire D, Tian C, Zhan X, Agee M, Alipanahi B, Auton A, Bell RK, Bryc K, Elson SL, Fontanillas P, Furlotte NA, Hinds DA, Hromatka BS, Huber KE, Kleinman A, Litterman NK, McIntyre MH, Mountain JL, Northover CAM, Sathirapongsasuti JF, Sazonova OV, Shelton JF, Shringarpure S, Tian C, Tung JY, Vacic V, Wilson CH, Pitts SJ, Mitchell A, Skogholt AH, Winsvold BS, Sivertsen B, Stordal E, Morken G, Kallestad H, Heuch I, Zwart J-A, Fjukstad KK, Pedersen LM, Gabrielsen ME, Johnsen MB, Skrove M, Indredavik MS, Drange OK, Bjerkeset O, Børte S, Stensland SØ, Choquet H, Docherty AR, Faul JD, Foerster JR, Fritsche LG, Gabrielsen ME, Gordon SD, Haessler J, Hottenga J-J, Huang H, Jang S-K, Jansen PR, Ling Y, Mägi R, Matoba N, McMahon G, Mulas A, Orrù V, Palviainen T, Pandit A, Reginsson GW, Skogholt AH, Smith JA, Taylor AE, Turman C, Willemsen G, Young H, Young KA, Zajac GJM, Zhao W, Zhou W, Bjornsdottir G, Boardman JD, Boehnke M, Boomsma DI, Chen C, Cucca F, Davies GE, Eaton CB, Ehringer MA, Esko T, Fiorillo E, Gillespie NA, Gudbjartsson DF, Haller T, Harris KM, Heath AC, Hewitt JK, Hickie IB, Hokanson JE, Hopfer CJ, Hunter DJ, Iacono WG, Johnson EO, Kamatani Y, Kardia SLR, Keller MC, Kellis M, Kooperberg C, Kraft P, Krauter KS, Laakso M, Lind PA, Loukola A, Lutz SM, Madden PAF, Martin NG, McGue M, McQueen MB, Medland SE, Metspalu A, Mohlke KL, Nielsen JB, Okada Y, Peters U, Polderman TJC, Posthuma D, Reiner AP, Rice JP, Rimm E, Rose RJ, Runarsdottir V, Stallings MC, Stančáková A, Stefansson H, Thai KK, Tindle HA, Tyrfingsson T, Wall TL, Weir DR, Weisner C, Whitfield JB, Winsvold BS, Yin J, Zuccolo L, Bierut LJ, Hveem K, Lee JJ, Munafò MR, Saccone NL, Willer CJ, Cornelis MC, David SP, Hinds DA, Jorgenson E, Kaprio J, Stitzel JA, Stefansson K, Thorgeirsson TE, Abecasis G, Liu DJ, Vrieze S, andMe Research T, Psychiatry HA-I: Association studies of up to 1.2 million individuals yield new insights into the genetic etiology of tobacco and alcohol use. Nature Genetics 2019;51:237-244

4. Burgess S, Thompson SG: Multivariable Mendelian Randomization: The Use of Pleiotropic Genetic Variants to Estimate Causal Effects. American Journal of Epidemiology 2015;181:251-260

5. Rees JMB, Wood AM, Burgess S: Extending the MR-Egger method for multivariable Mendelian randomization to correct for both measured and unmeasured pleiotropy. Statistics in Medicine 2017;36:4705-4718

6. R Core Team: R: A language and environment for statistical computing. Vienna, Austria, R Foundation for Statistical Computing, 2021

7. Hemani G, Tilling K, Davey Smith G: Orienting the causal relationship between imprecisely measured traits using GWAS summary data. PLOS Genetics 2017;13:e1007081

8. Hemani G, Zheng J, Elsworth B, Wade KH, Haberland V, Baird D, Laurin C, Burgess S, Bowden J, Langdon R, Tan VY, Yarmolinsky J, Shihab HA, Timpson NJ, Evans DM, Relton C, Martin RM, Davey Smith G, Gaunt TR, Haycock PC: The MR-Base platform supports systematic causal inference across the human phenome. eLife 2018;7:e34408

9. Sanderson E, Spiller W, Bowden J: Testing and correcting for weak and pleiotropic instruments in two-sample multivariable Mendelian randomization. Statistics in Medicine 2021;40:5434-5452
